# Improving outcomes for people who are homeless and have severe mental illness in Ethiopia, Ghana and Kenya: overview of the HOPE programme

**DOI:** 10.1101/2024.11.28.24318066

**Authors:** Charlotte Hanlon, Caroline Smartt, Victoria Mutiso, Peter Yaro, Eleni Misganaw, Ursula Read, Rosie Mayston, Ribka Birhanu, Phyllis Dako-Gyeke, David Ndetei, Laura Asher, Julie Repper, Julian Eaton, Kia-Chong Chua, Abebaw Fekadu, Ruth Tsigebrhan, Cecilia Ashaley Fofo, Kimberly Kariuki, Sauharda Rai, Sisay Abayneh, Caroline Reindorf Amissah, Amma Mpomaa Boadu, Priscilla Makau, Agitu Tadesse, Phil Timms, Martin Prince, Graham Thornicroft, Brandon Kohrt, Atalay Alem

**Affiliations:** Division of Psychiatry, Centre for Clinical Brain Sciences, University of Edinburgh, Edinburgh, Scotland, UK; and Centre for Innovative Drug Development and Therapeutic Trials for Africa (CDT-Africa), College of Health Sciences, Addis Ababa University, Addis Ababa, Ethiopia.; Centre for Global Mental Health, Health Service and Population Research Department, Institute of Psychiatry, Psychology and Neuroscience, King’s College London, London, UK.; Africa Institute of Mental and Brain Health, Nairobi, Kenya. email; BasicNeeds-Ghana, Tamale, Ghana.; Mental Health Service User Association, Addis Ababa, Ethiopia; School of Health and Social Care, University of Essex, Colchester, Essex, UK; Department of Global Health & Social Medicine, King’s College London, UK.; Addis Ababa University, College of Health Sciences, School of Medicine, Department of Psychiatry and WHO Collaborating Centre in Mental Health Research and Capacity-Building.; Department of Social and Behavioural Sciences, School of Public Health, University of Ghana.; Professor of Psychiatry, University of Nairobi and Africa Institute of Mental and Brain Health, Nairobi, Kenya.; Centre for Public Health and Epidemiology, School of Medicine,□University of Nottingham, Nottingham, UK; Institute of Mental Health, University of Nottingham, Nottingham, UK; Imroc (Charity Reg No 1207904), Nottingham, UK; CBM Global and Liverpool School of Tropical Medicine, Liverpool UK; Institute of Psychiatry, Psychology, and Neuroscience, King’s College London.; Centre for Innovative Drug Development and Therapeutic Trials for Africa (CDT-Africa), College of Health Sciences, Addis Ababa University, Addis Ababa, Ethiopia; and Department of Global Health & Infection, Brighton and Sussex Medical School, Brighton, UK.; Addis Ababa University, College of Health Sciences, School of Medicine, Department of Psychiatry and WHO Collaborating Centre for Mental Health Research and Capacity-Building. Email-; BasicNeeds-Ghana, Accra, Ghana.; Kenya Medical Research Institute - Centre for Clinical Research, Division of Mental Health; Center for Global Mental Health Equity, Department of Psychiatry and Behavioral Health, George Washington University, Washington, DC, USA.; Arsi University, College of Education and Behavoural Studies, Arsi Asela, Ethiopia; and Department of Psychiatry, School of Medicine, College of Health Sciences, Addis Ababa University, Addis Ababa, Ethiopia; Deputy Chief Executive, Mental Health Authority, Accra, Ghana.; Mental Health Department, Institutional Care Division, Accra, Ghana Health Service.; Kitui County, Ministry of Health, Nairobi, Kenya; Federal Ministry of Health of Ethiopia, Addis Ababa, Ethiopia; National Psychosis Unit, London, UK; and King’s College London, UK. Email: South London and Maudsley NHS Foundation Trust.uk; Department of Public Health Sciences, Faculty of Life Sciences and Medicine, King’s College London, UK.; Centre for Global Mental Health and Centre for Implementation Science, Health Service and Population Research Department, Institute of Psychiatry, Psychology & Neuroscience, King’s College London, London, UK.; Center for Global Mental Health Equity, Department of Psychiatry and Behavioral Health, George Washington University, Washington, DC, USA; Department of Psychiatry and WHO Collaborating Centre in Mental Health Research and Capacity-Building, School of Medicine, College of Health Sciences, Addis Ababa University, Addis Ababa, Ethiopia.

**Keywords:** Africa, community mental health, intervention, homelessness, participatory action research, psychosis, severe mental illness, serious mental illness

## Abstract

**Aim:** HOPE (National Institute for Health and Care Research Global Health Research Group on Homelessness and Mental Health in Africa) aims to develop and evaluate interventions that address the unmet needs of people who are homeless and have severe mental illness (SMI) living in three African countries in ways that are rights-based, contextually grounded, scalable and sustainable.

**Methods:** We will work in the capital city (Addis Ababa) in Ethiopia, a regional city (Tamale) in Ghana, and the capital city (Nairobi) and a rural county (Makueni) in Kenya to understand different approaches to intervention needed across varied settings.

Formative work will include synthesis of global evidence (systematic review, including grey literature, and a Delphi consensus exercise) on interventions and approaches to improving outcomes for people who are homeless and have SMI. We will map contexts; conduct a focused ethnographic study to understand lived experiences of homelessness and SMI; carry out a cross-sectional survey of people who are homeless (n=750 in Ghana and Ethiopia; n=350 in Kenya) to estimate prevalence of SMI and identify prioritised needs; and conduct in-depth interviews and focus group discussions with key stakeholders to understand experiences, challenges and opportunities for intervention. This global and local evidence will feed into Theory of Change workshops with stakeholders to establish agreement about valued primary outcomes, map pathways to impact and inform selection and implementation of interventions. Intervention packages to address prioritised needs will be co-produced, piloted in each country with people who are homeless and have SMI, and will be optimised for feasibility and acceptability using participatory action research. We will use rights-based approaches and focus on community-based care to ensure sustainability. Realist approaches will be employed to analyse how contextual variation affects mechanisms and outcomes to inform methods for a subsequent evaluation of larger scale implementation. Extensive capacity strengthening activities will focus on equipping early career researchers and peer researchers. People with lived experience of SMI and policymakers are an integral part of the research team. Community engagement is supported by working closely with multi-sectoral Community Advisory Groups.

**Conclusions:** HOPE will develop evidence to support action to respond to the needs and preferences of people who are homeless and have SMI in diverse settings in Africa. We are creating a new partnership of researchers, policy makers, community members and people with lived experience of SMI and homelessness to lead this work in and for the Global South.

## Background

People with severe mental illness (SMI; comprising disabling psychoses and affective conditions (often co-morbid with substance use problems) are over-represented in homeless populations globally. However, the situation is stark in low- and lower-middle-income countries (LLMICs), where an estimated 28-36 million people are homeless and have SMI (Chamie 2017; Smartt et al. 2019). In systematic reviews from high-income countries, the co-occurrence of SMI and homelessness is associated with numerous adverse outcomes, including premature mortality (Fazel et al. 2014); infectious disease (chiefly tuberculosis, HIV and hepatitis B)(Beijer et al. 2012); non-communicable diseases (Scott et al. 2013); co-morbid alcohol and substance abuse; injuries and accidents (Mackelprang et al. 2014); and suicide (Arnautovska et al. 2014). Women and youth who are homeless are at particular risk of sexual assault and exploitation (Goodman et al. 1995). SMI is a major risk factor for chronic homelessness (Fazel et al. 2014).

Despite strong social capital and protective family structures in many LLMICs in Africa, caring for a person with SMI can overwhelm informal support networks, exacerbated by precarious household finances, stigma, diverse understandings about mental illness and limited mental healthcare provision (Read et al. 2009). Harsh conditions on the streets can also trigger new onset of SMI. High quality evidence from LLMICs is scarce, but most people who are homeless and have SMI have unmet basic needs for water, clothing and food, 75% with untreated medical problems, and 30-40% affected by physical disabilities (Fekadu A et al. 2014; Singh et al. 2016; Tripathi et al. 2013). Beyond basic needs, there is also evidence of high levels of unmet needs for social support, relationships and rehabilitation (80%), and of exposure to unjust imprisonment, exploitation, physical and sexual abuse. Few (10%) have ever received mental healthcare, and almost none received adequate ongoing care.

The evidence base for intervention in high-income countries takes housing as an essential aspect of service provision for people who have SMI and are homeless, particularly the Housing First model (Tsemberis 2011), which improves a wide variety of health and social outcomes (Woodhall-Melnik and Dunn 2016). Other evidence-based health interventions include tailored primary healthcare programmes, care co-ordination, assertive community mental health treatment and critical time interventions (Hwang and Burns 2014). The applicability of such models to LLMICs is likely to be limited, given their reliance on access to housing, specialist multi-disciplinary workers and government-funded social welfare and healthcare services. Furthermore, factors causing or maintaining homelessness in people with SMI in LLMICs may vary and demand different responses, for example, in relation to mental health stigma and discrimination, forced displacement and poverty.

In our scoping review (Smartt et al. 2019), we were unable to identify any rigorous evaluations of programmes for people who are homeless and have SMI in LLMICs. Common components of identified programmes included: residential care; mental healthcare and vocational/occupational training. Most programmes are run by stand-alone, non-governmental organisations (NGOs), rarely integrated with government provision (Narasimhan et al. 2019), thus limiting scalability and sustainability. Crucially, little attention is paid to social needs and the preferences of those with lived experience, who may not wish to return to their communities of origins, due to ’push’ factors such as shame, a need to escape from conflict, broken relationships, exploitative work, or other forms of abuse and exclusion or because of ‘pull’ factors related to perceived opportunities and freedoms in cities (Haile et al. 2020).

There remains a crucial gap in evidence and global recommendations of care for people with SMI who are homeless. Any effective response will require co-ordination and close collaboration of specialist and community-based care across sectors, particularly when family supports are overwhelmed (Hanlon 2017) or absent, as well as the meaningful involvement of people with lived experience. This evidence gap indicates a pressing need for an Africa-led partnership of researchers and key stakeholders to conduct high quality research in this area.

## Aims and Objectives

The aim of HOPE (National Institute for Health and Care Research Global Health Research Group on Homelessness and Mental Health in Africa) is to develop and evaluate approaches to addressing the unmet needs of people who are homeless and have SMI living in LLMICs in ways that are rights-based, contextually grounded, scalable and sustainable.

Objectives of HOPE:

1. Establish a partnership of researchers, implementers, policymakers and people with lived experience.
2. Synthesise global evidence and obtain expert consensus on priority actions and approaches.
3. Work in distinctive settings in Ethiopia (capital city), Ghana (regional capital) and Kenya (capital city and rural county) to:

a. Identify the priority needs and valued outcomes of those with lived experience, and opportunities and challenges for intervention.
b. Integrate global and local evidence to select, co-produce and pilot interventions that target priority needs, in order to investigate acceptability and feasibility.
c. Evaluate the impact of interventions on the human rights and outcomes valued by people with lived experience and generate evidence on intervention costs, implementation processes and outcomes.
d. Pioneer the development of methods and ethical frameworks for future research and interventions in this area.
4. Impact global policy and practice through translation of evidence into a ‘how-to’ guide to adapt and implement programmes in diverse LLMIC contexts.
5. Build sustainable capacity across partners that responds to local priorities, enhances existing capabilities, supports South-South and South-North exchange of expertise and mutual learning, and develops individuals, teams, organisations and systems.

## Methods

See Figure 1 for an overview of HOPE. HOPE is structured into work packages (WP): WP1 (project co-ordination), WP2 (formative phase), WP3 (intervention co-production/piloting), WP4 (implementation/evaluation), WP5 (capacity-strengthening) and WP6 (lived experience, community engagement and research uptake). In HOPE, we have prioritised the involvement of people with lived experience in all aspects of the research and the delivery of interventions.

**Figure 1:**
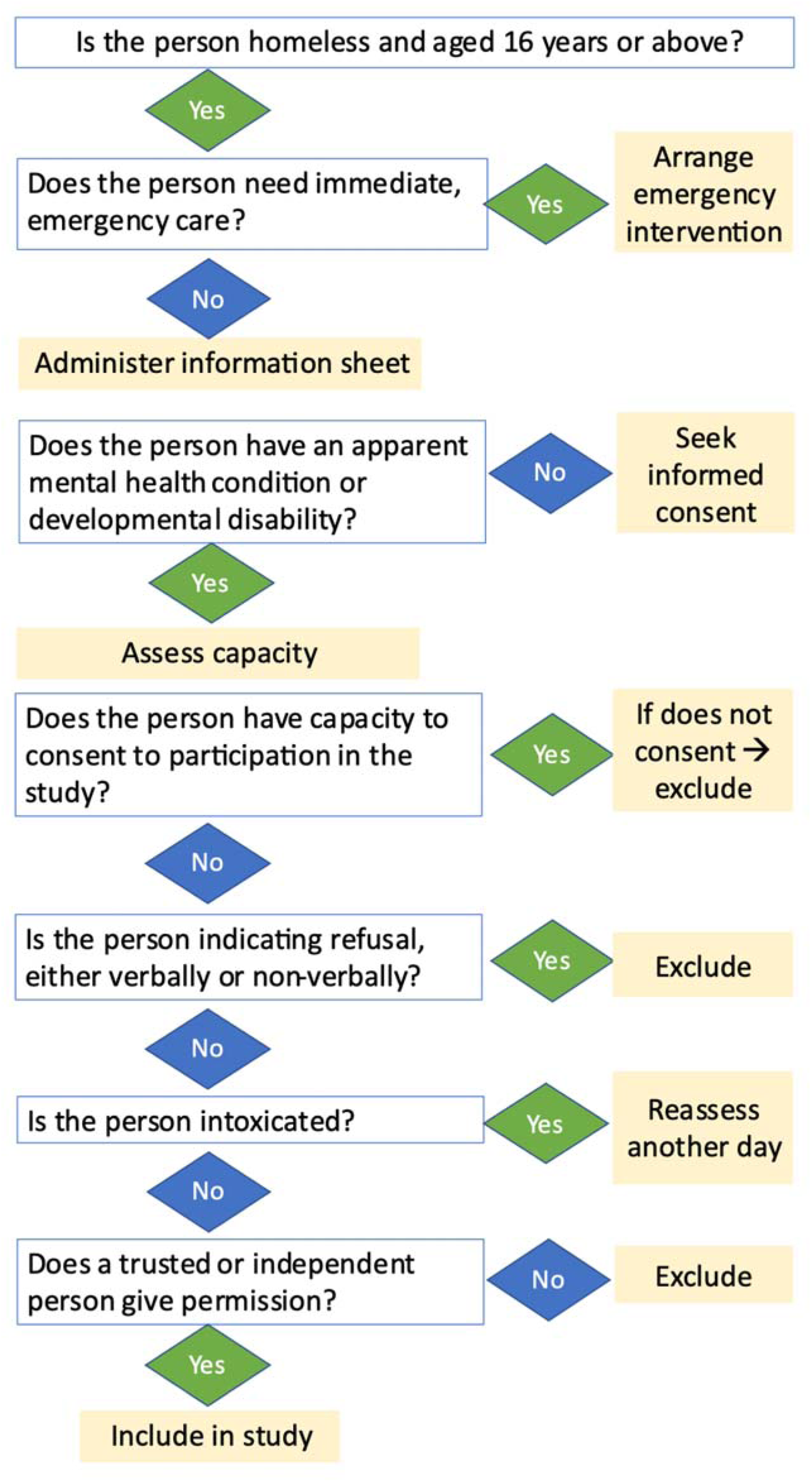
Overview of procedures for recruitment of a person with severe MHC who lack capacity to consent – Part 1

**Figure 2:**
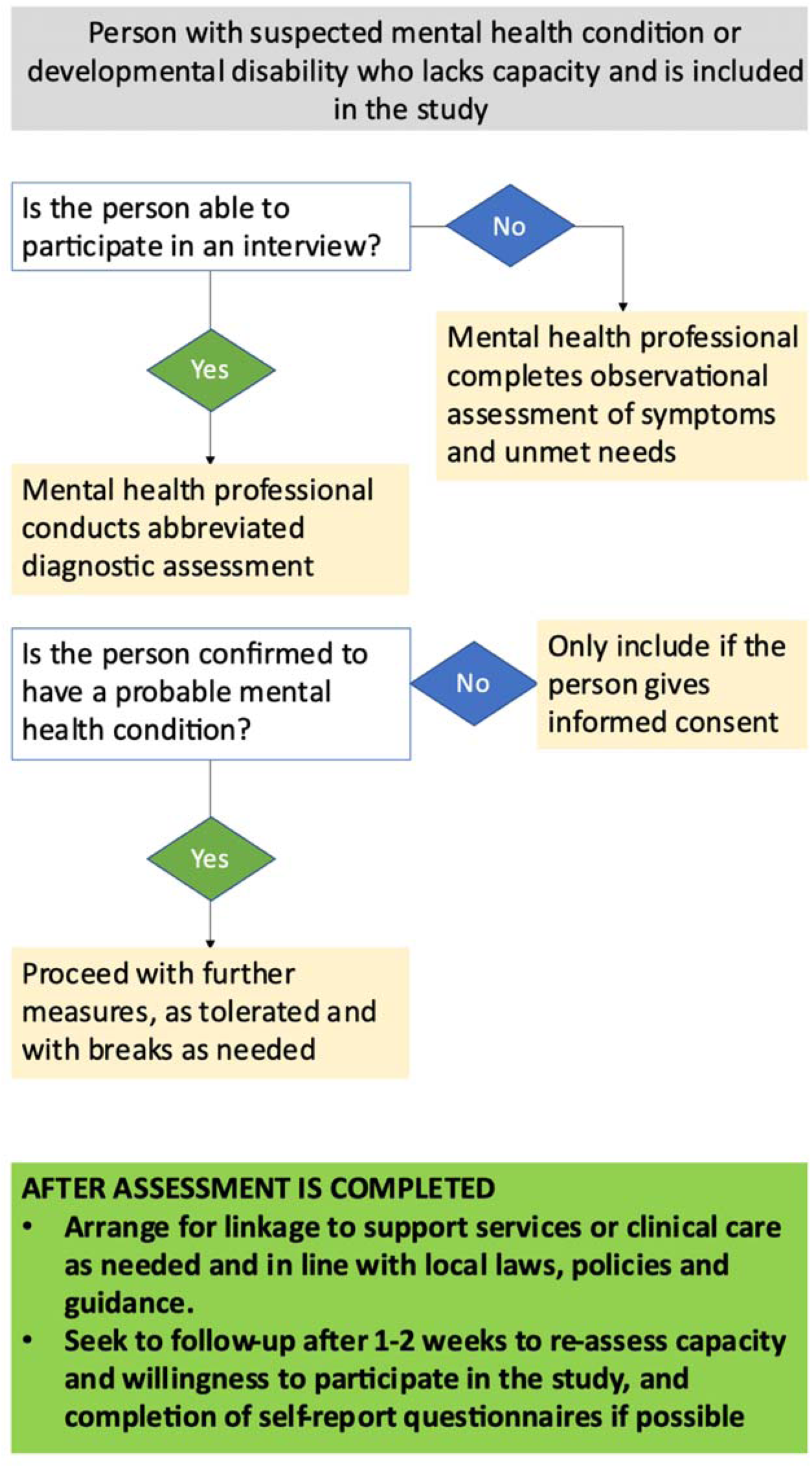
Overview of procedures for recruitment of a person with severe MHC who lack capacity to consent – Part 2

## Countries and settings

See Table 1 for characteristics of the countries and sites. These sites have been selected because: (1) there is an evident need in each setting to improve support for people who are homeless and have SMI, (2) they are distinctive contexts in East and West Africa across low-income and lower-middle income countries; and (3) there are opportunities to build on existing programmes and political will. Homelessness has increased across the study countries, with accompanying challenges of poor health outcomes (Elsey et al. 2019). There are no reliable estimates of how many people are homeless in the study countries. In each study setting, social exclusion and stigmatization of people who are homeless is common (UNICEF et al. 2019), which intersects with stigma and discrimination experienced by persons with SMI (Forthal et al. 2019; Mutiso V. N. et al. 2019; Read et al. 2009).

**Table 1:** Characteristics of countries and project sites in HOPE

|  | Ethiopia | Ghana | Kenya |  |
| --- | --- | --- | --- | --- |
| Country location | East Africa | West Africa | East Africa |  |
| World Bank income category | Low-income | Lower-middle | Lower-middle |  |
| Country population (World Bank Group 2023) | 126 million | 34.1 million | 55.1 million |  |
| Gross Domestic Product/capita (World Bank Group 2023) | \$1294 | \$2238 | \$1950 | |
| Number of mental health professionals/100,000 population | 0.11 psychiatrists, 0.58 master’s level practitioners/psychiatric nurses, 0.045 clinical psychologists and 0.009 social workers (Federal Democratic Republic of Ethiopia Ministry of Health 2021) | 0.07/ 100,000 (Roberts et al. 2014) | 0.19/100,000 (WHO 2014) |  |
| Evidence on burden of homelessness and SMI | 41% prevalence of SMI in sample of 217 people who were homeless in Addis Ababa, with extensive unmet needs (Fekadu et al. 2014) | No prevalence estimates. Research has mostly focused on homeless youth in Accra (Asante et al. 2014). | No data available |  |
| Previous experience with interventions for people who are homeless and have SMI | Linked to expanding access to mental health care in a rural district in south-central Ethiopia (Smartt et al. 2021) | In Tamale, community mental health workers assess and treat people who are homeless and have SMI within their catchment area. | In Nairobi, the CREATE initiative (Community Recovery Achieved Through Entrepreneurism) demonstrated how an integrated approach to social inclusion of people with SMI, including some who were homeless (Casey et al. 2018) could support employment of people with SMI (MacDougall et al. 2021) |  |
| HOPE project sites |  |  |  |  |
|  | Ethiopia | Ghana | Kenya |  |
| HOPE project sites | Addis Ababa sub-cities (Arada, Addis Ketema) | Tamale city | Nairobi county (Kamukunji, Starehe, Embakasi sub-counties) | Makueni county |
| Population of project site | 4.4 million for Addis Ababa (United Nations Department of Economic and Social Affairs Population Division (2018) – with minor processing by Our World in Data 2018) | 375,000 (Tamale Metropolitan Authority 2021) | 4.4 million (United Nations Department of Economic and Social Affairs Population Division (2018) – with minor processing by Our World in Data 2018) | 1 million (Ngugi 2013) |
| Characteristics of project site | These sub-cities have lower SES as well as concentrated numbers of people who are homeless compared with the other sub-cities in Addis Ababa. They | Rapidly expanding regional capital situated 620 km from the capital; relatively lower | Nairobi occupies an area of about 700 km<sup>2</sup> at the south-eastern end of Kenya’s agricultural | 250 km southeast of Nairobi, lower SES, with over 60% of the population |
|  | also feature transport hubs as well as the main market in Addis Ketema. Overcrowding and poor quality of housing is a feature of both sub-cities, although new government-led housing projects have been constructed in the past 5-10 years. | socio-economic status in Ghana; situated on major internal and international transport routes; regional trading and agricultural centre | heartland. Nairobi is facing challenges of rapid urbanization, which is accompanied by informal settlements with poor quality of housing, as well as air and water pollution, water supply and sanitation. | living below the poverty line (Government of Makueni County 2018). |
| Mental health services | 443 in-patient beds; hospital-based out-patient services; some integration in primary care; 1 residential rehabilitation facility | Hospital-based in-patient and out-patient services; community outreach by mental health nurses | Partially integrated mental health services at primary health care levels | Integrated mental health services at the primary healthcare level (Mutiso V. N. et al. 2019) |
| Social care | Limited provision to meet basic needs from NGOs and religious institutions | NGOs and civil society organizations provide ad hoc support | Provision of social services is provided in a fragmented manner, either by faith-based organizations or NGO and other civil society organizations. There is a draft policy on street families that is yet to be finalized but no current services for people who are homeless. | No mental health or other public sector services for people who are homeless |

Although all three countries are expanding community-based mental healthcare, these services are not tailored to the complex needs of people who are homeless and are inaccessible due to the reliance on families to bring a person to services and often meet the costs of treatment. Faith-based, civil society and NGOs contribute to crisis responses to basic needs for sub-groups of homeless populations but such efforts are fragmented (UNICEF et al. 2019).

### Conceptualising homelessness

Our starting point for defining homelessness is: spending the night unsheltered or in other places not intended for habitation. We will exclude people who spend their days on the streets, for example, to beg, but who have stable night-time accommodation. However, the concept of homelessness is complex and locally nuanced, so the target population will be operationalised for each study site in the formative phase.

### Theoretical framework

The research work in HOPE will follow the Medical Research Council/National Institute for Health and Care Research framework for developing and evaluating complex interventions (Skivington et al. 2021). We will use Theory of Change (ToC) as a participatory tool to inform each aspect of the MRC approach (De Silva et al. 2014). Applying realist approaches, we will develop programme theories to inform evaluation. Unmet needs will be framed within a socio-ecological model that seeks to understand and propose interventions to address the individual, family, societal and political-level barriers to social inclusion.

### Work Package 2: Formative Phase

An extensive formative phase will comprise syntheses of global evidence and best practice alongside primary data collection in each setting: context mapping, ethnography and a cross-sectional study.

#### (i) Systematic review

Building on our previous scoping review (Smartt et al. 2019), we will conduct a systematic review, including grey literature, to identify interventions for people who are homeless with SMI in LLMICs, as per the published protocol (Smartt et al. 2023). We will carry out a narrative synthesis of types of interventions, implementation strategies, and evidence of impact.

#### (ii) Delphi consensus exercise

A Delphi consensus exercise will help to identify global perspectives on best practices and priorities for interventions. We will invite people with experience of implementing relevant programmes in different regions in LLMICs, and representatives of international mental health service user associations, disability rights organisations and the World Health Organization (WHO). In the first round, participants identify interventions/ components of programmes that they consider important. These will be consolidated and augmented by emerging findings from our review. In the second round, participants will rank each identified component/strategy based on importance and feasibility in LLMICs. Finally, the anonymised aggregated responses will be fed back to the participants, with further ranking to identify essential, desirable and optional elements of intervention programmes, as well as any ethical concerns.

#### (iii) Context mapping

We will conduct a documentary analysis of relevant policies, laws and plans relating to people who are homeless in our settings, inclusive of documents relating to public health, social care, legal decrees, and other municipal documents. We will consult with community leaders and other key stakeholders in administrative positions, NGOs and religious and traditional healing sites. We will establish where people who are homeless and have SMI can be reached, document the roles of different agencies and existing initiatives. Findings from the analysis will be synthesised using a matrix, informed by an implementation framework to map context (Pfadenhauer et al. 2017), facilitating cross-country comparisons.

#### (iv) Primary data collection for formative studies

The formative research involving primary data collection comprises a focused ethnography (Sangaramoorthy and Kroeger 2020) and a cross-sectional study to understand experiences of homelessness and SMI, population burden, level of unmet needs, and sources of support, preferences, opportunities and potential barriers to intervention. We will carry out participant observation in locations where people are likely to sleep, visit or spend the day. Observations and linked interviews will focus on people who are homeless and have SMI, other members of the community, and their interactions. Interviews and focus group discussions will be conducted with key informants. The specific aims and methods are detailed in Table 2.

**Table 2:**
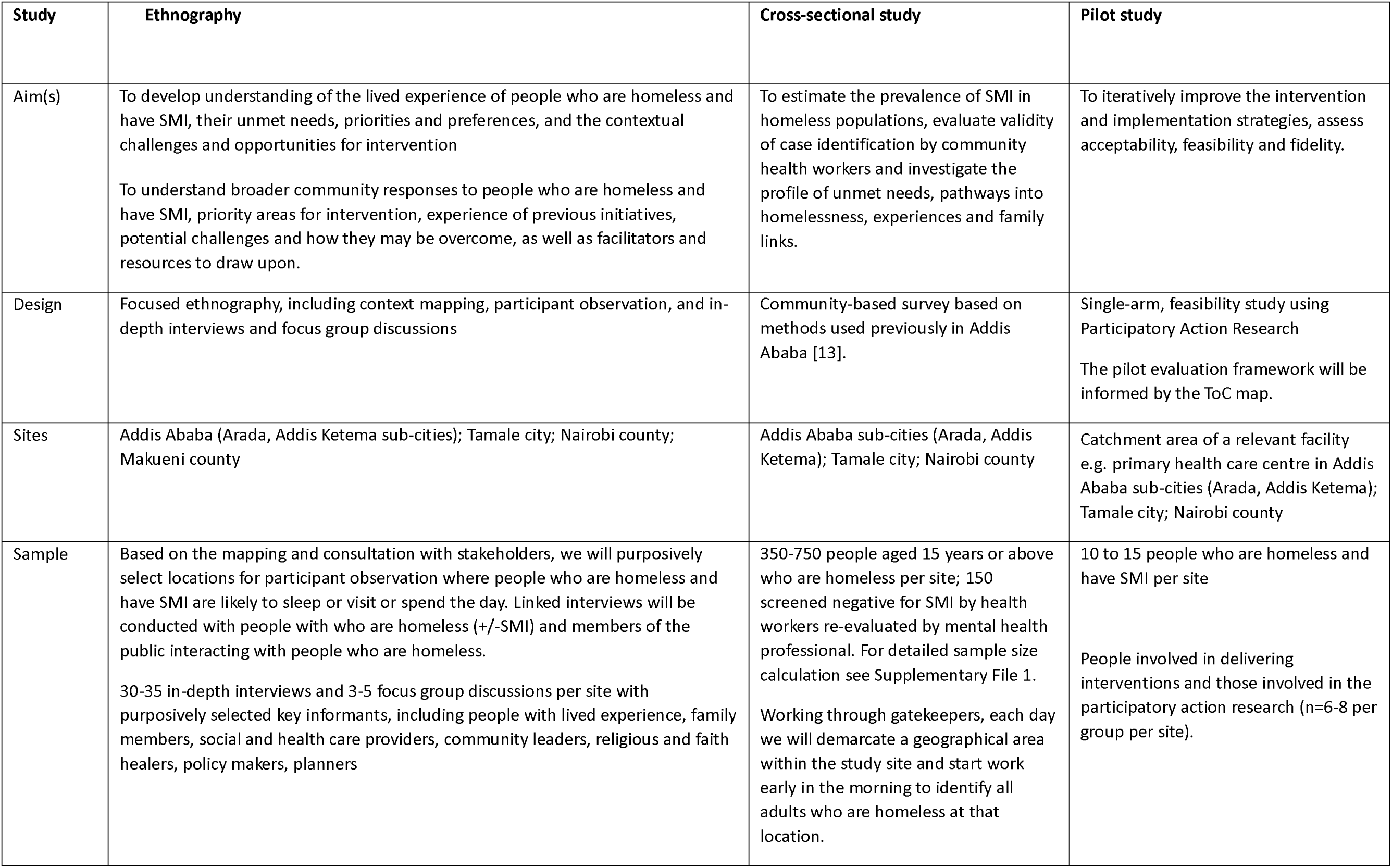
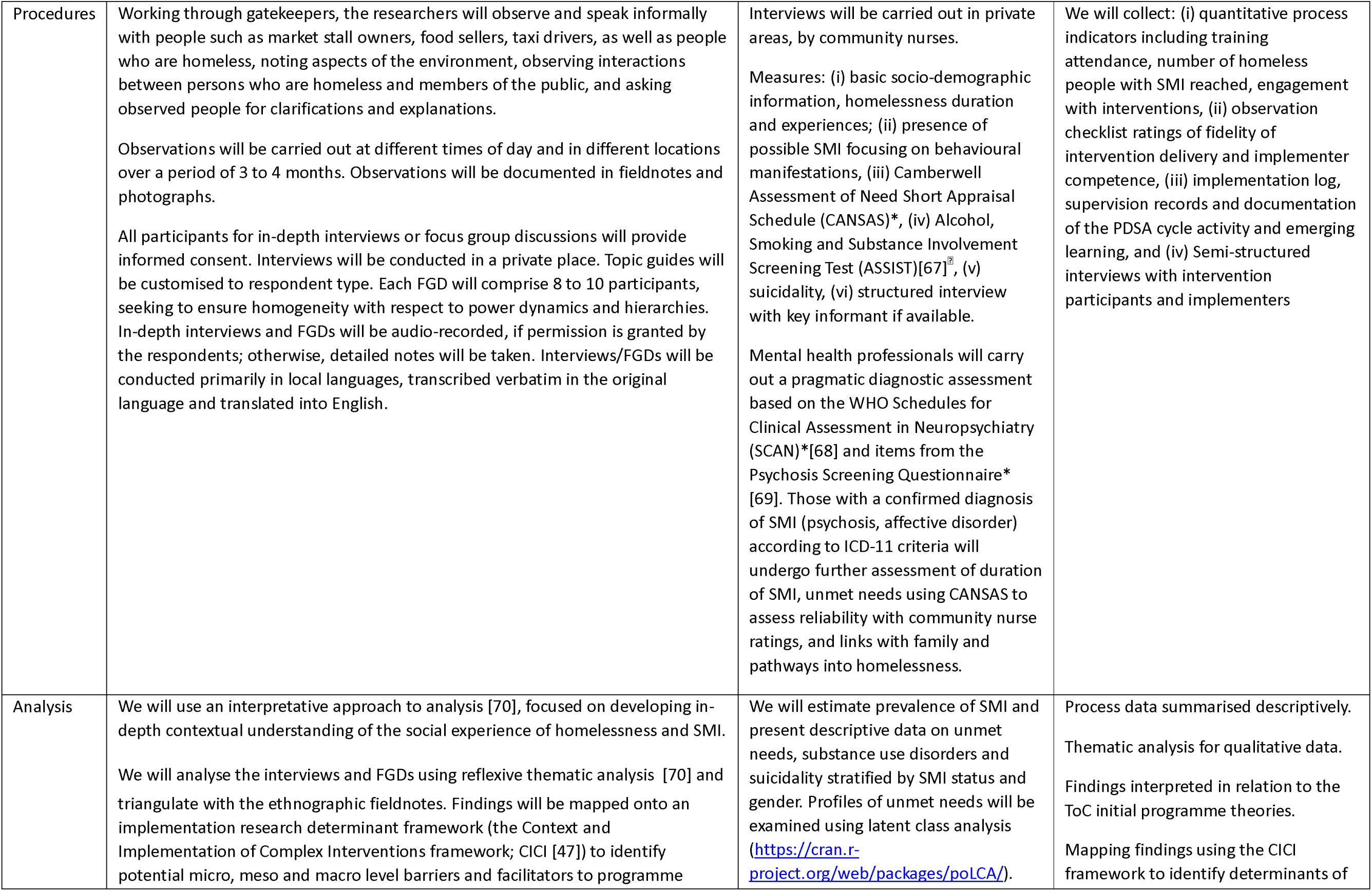
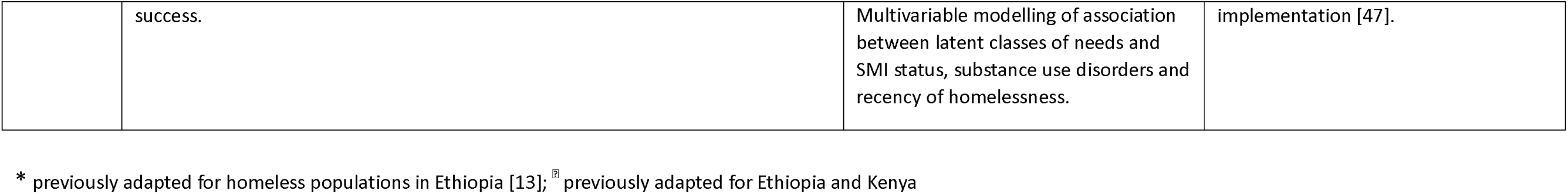
Overview of methods for formative phase studies and pilot study

### Ethics

Key ethical challenges relating to these studies will now be highlighted: consent, responding to basic needs and safeguarding.

#### Consent to participate

In the ethnography, gatekeeper permission will be obtained. Aligned with accepted ethics of ethnographic practice, we will not seek formal consent from individuals who are being observed. The purpose of participant observation is to access an understanding of the day-to-day lives of people who are homeless and have SMI which might not be obtained via other methods, such as formal qualitative interviews. The process of obtaining informed consent can disrupt participants’ natural behaviours, actions and responses, including their relationship with the researcher-observer, risking the valuable insights ethnography might add. This was a point of contention in the HOPE team, with concerns raised by the Lived Experience Advisory Group (below) about the dignity and autonomy of a person who is homeless and has SMI. To address these concerns, we will seek to build rapport with people who are homeless and have SMI over time through regular visits by the researchers and informal conversation, an approach used to engage homeless people with SMI in similar settings (Eaton et al. 2015). This will facilitate respectful engagement with the person, allowing them to gain an understanding of the researcher’s role and develop trust. The researchers will be carefully trained and supervised to ensure they respect any desire not to be observed and cease observation if a person communicates discomfort (verbally or non-verbally). For in-depth interviews, informed consent will be obtained.

In the cross-sectional study, we will include people who lack mental capacity to provide informed consent as participants in the study with appropriate safeguards. We argue that the ethical principle of justice is upheld because the study enables us to obtain an understanding of the needs of the most vulnerable people who are homeless and have SMI and design interventions that best meet those needs. The United Nations Convention on the Rights of Persons with Disabilities implies a presumption of equal treatment with others who may wish to participate in research (i.e. having equal legal capacity), with the proviso that the prospective participant’s will and preference is always actively recognised and respected. We will not include anyone in the study if they indicate refusal or if there is any other indication that their will and preference would be not to participate. To identify evidence of the latter, we will seek to speak to a trusted person who is nominated by the person who is homeless and has SMI. That person could be anyone who credibly supports the individual. In the absence of such a person, a mental health/disability advocate will communicate with the person, seek to identify their will and preference, and provide permission for the person’s participation.

Mental capacity will be assessed by a mental health professional, using a standardised approach (Hanlon et al. 2016). As a further safeguard for study participants who lack mental capacity, we will make systematic efforts to re-assess capacity after two weeks, during which time we will support them to access mental health services. If the person has regained capacity, they will be invited to provide informed consent to participate and, if they decline, they will be withdrawn from the study.

#### Responding to basic needs

During the ethnography fieldwork, researchers may offer refreshments to people who are homeless during informal interactions as an act of reciprocity and following altruistic norms. For the cross-sectional survey, refreshments will be made available for all people who are homeless in the vicinity, regardless of study participation, so as not to provide undue incentives or coercion to participate.

People with emergency medical needs will be supported to access care, working with local healthcare services and community health workers. When safeguarding needs surface, we will respond as detailed below. Participants will be informed about resources in the vicinity where they may access support e.g. in relation to shelter, feeding programmes, local health services, and support for vulnerable women. For people with SMI, concerted efforts will be made to link them to mental health services (according to their preference). Involuntary mental healthcare will be utilised only in line with the country’s legislation (Ghana, Kenya) or, where no legislation exists (Ethiopia), if the person’s mental state is assessed as posing an imminent risk of harm to themselves or others. In the previous study from Ethiopia (Fekadu et al. 2014), only two out of 89 people who were homeless and had SMI received involuntary psychiatric treatment.

#### Safeguarding

Standard operating procedures (See supplementary file 2]) have been developed to support researcher responses to safeguarding concerns arising from: (1) chaining, restraint or seclusion; (2) sexual abuse, exploitation or harassment; (3) physical abuse or serious physical health concerns; (4) suicidal behaviour; (5) violent or aggressive behaviour; (6) human rights abuses by professionals or community members; and (7) child trafficking or other child protection needs. Community advisory boards informed the development of localised protocols.

### Work Package 3: Intervention co-development and piloting

#### (i) Participatory theory of change workshops and intervention selection

We will conduct theory of change (ToC) workshops at the beginning and end of the formative phase. Each ToC workshop will last 0.5-1 day. Participants will include stakeholders identified through the formative work. Workshops will start by developing agreement on the desired long-term outcomes and impacts of the programme, then map out the interventions needed to achieve intermediate outcomes, identifying underlying assumptions and barriers, potential implementation strategies, required resources and inputs, and indicators of success. Global and country-specific evidence and experience arising from the formative phase findings will be integrated into the initial programme theory derived from the ToC workshops.

Interventions with evidence of effectiveness in low- or middle-income countries (identified through our reviews and Delphi exercise) will be examined for relevance to preferences, unmet needs and feasibility of adaptation (based on formative studies). We will identify where new intervention development is needed (Box 1). This process will initially be undertaken within each country by a small (n=5-8) working group comprising people with lived experience of SMI, potential implementers and the HOPE country project teams. HOPE consortium members will then review the proposed interventions/approaches and suggest further ways to tailor the interventions to the local context.

We will give particular focus to rights-based approaches, potential scalability and sustainability, and identifying the role of peer support.

#### (ii) Co-development of interventions and implementation strategies

Working groups of people with lived experience, implementers and the research team will meet several times to co-produce the interventions, implementation strategies and fidelity checklists. We will describe the interventions according to recommended guidance (Hoffmann et al. 2014)) and catalogue the implementation strategies according to existing taxonomies (Powell et al. 2015) to enhance testing in new settings. The FRAME implementation tool (Framework for Reporting Adaptations and Modifications Expanded) (Wiltsey Stirman et al. 2019) will be used to document intervention adaptations and their justification. We anticipate that the resulting intervention packages for the three study settings will have elements that are similar, although addressed in differing ways depending on the context (e.g. accessing physical and mental healthcare, addressing basic needs, common elements of needs assessment and planning, peer support, individual engagement, addressing social exclusion) as well as elements that may be site-specific (e.g. addressing substance misuse, family interventions, housing). Candidate personnel for delivering and supervising the intervention/s will be existing human resources, such as community health workers, peers (people with lived experience), the NGO sector, social workers, primary healthcare staff and psychiatric nurses.

#### (iii) Realist methods

We will conduct a realist synthesis to inform our understanding of what works, for whom, in what circumstances and why (Wong et al. 2017). Drawing on cross-country analyses of findings from the formative phase, we will map our initial ToC-based programme theories onto mid-range programme theories and, in collaboration with key stakeholders, develop hypotheses about the important ways that interventions can bring about change in different contexts i.e. ‘context-mechanism-outcome (CMO) configurations’. These will then be explored in the pilot phase and used to develop a realist evaluation framework for larger-scale implementation.

#### (iv) Pilot study

The resulting intervention package will be piloted in each country in a circumscribed geographical area linked to a primary healthcare centre or other relevant facility. We will conduct a single arm feasibility study over three months, focusing on initial assessment and care planning, engagement and early interventions. See Table 2 for detailed methods. Based on the ToC and realist synthesis, we will identify process indicators (spanning quantitative and qualitative data) for each intermediate outcome and explore potential mechanisms through which context influences outcomes. We will examine the feasibility and appropriateness of using routinely collected service indicators and identify where project data will be needed as the basis for monitoring and evaluation, and quality improvement. We will initially run training for a small group of implementers. Regular meetings (every 1-2 weeks) with implementers and people with lived experience, supported by the project team, will use plan-do-study-act (PDSA) cycles to identify and address emerging challenges. The structures and supports that underpin this co-production approach are the focus of work packages 5 and 6.

#### (v) Developing tools to cost the interventions

Proformas will be designed and tested to facilitate key decisions about our costing approach; for example, determining which intervention components can be reliably estimated at an individual (bottom-up) level and which will require an aggregate (top-down) approach. In semi-structured interviews with implementers, we will gain views on the extent to which additional tasks related to the intervention replace or add to existing activities, to identify possible resource allocation for services or organisational implications that need to be scrutinised in the larger-scale implementation and evaluation study. A service use data collection form will be adapted and piloted.

### Work Package 4: Implementation and evaluation

Based on the pilot study findings, the country working groups will finalise interventions and implementation strategies. We will then carry out phased implementation of the co-produced and piloted intervention package at a larger scale in the formative study sites. A study protocol for WP4 will be reported separately.

### Work Package 5: Capacity strengthening

We are not aware of a partnership focused on addressing needs of people who are homeless and have SMI in Africa. Tackling this problem mandates use of research methodologies that have not been widely used in global mental health, including ethnography and participatory action research. To be successful, the partnership also needs to expand research capabilities to peer researchers and implementers, build researcher capacity for rights-based work and strengthen capacity for multi-sectoral working that is respectful and inclusive. We will deliver a programme of capacity-strengthening that builds on identified capabilities, is tailored to the different needs across countries and institutions, links directly to the work in HOPE, is designed for sustainability and is evaluated for impact (Hanlon et al. 2018). See Table 3 for capacity strengthening activities in HOPE. Guided by the ESSENCE framework (TDR/World Health Organization 2016) and our previous approaches, our capacity strengthening efforts will be evaluated for impact by collecting the following data: process indicators (participants in training courses and webinars: gender, age and background of participants; uptake and completion of training), satisfaction (anonymous online survey), impact (publications, percentage of first authors from LMICs, with lived experience and/or who are female, grant applications and conference presentations).

#### Box 1

Candidate interventions for adaptation

HOPE co-investigators have been involved in the development, implementation and evaluation of several interventions for people with SMI in the study countries which have potential relevance to homeless populations. Materials (manuals, training materials) and implementation strategies (e.g. for competency-based training and supervision (Laura Asher et al. 2021)) are available for the following: community-based rehabilitation for people with SMI delivered by lay workers (Asher et al. 2022; Asher et al. 2018), social contact-based stigma reduction interventions (Kohrt et al. 2021), awareness-raising and community outreach for people who are homeless and have SMI, engagement with traditional and religious healers (L. Asher et al. 2021; Yaro et al. 2020), task-shared models of integrated mental health care in primary health care (Hanlon et al. 2022; Hanlon et al. 2019; Mutiso V. N. et al. 2019), brief psychological interventions (Bitew et al. 2021), brief interventions for substance use disorders (Clair et al. 2019; Hailemariam et al. 2016; Harder et al. 2020) and gender-based violence (Keynejad et al. 2020), family interventions (Asher et al. 2018; Casey et al. 2018), livelihoods interventions (BasicNeeds-Ghana ; Lund et al. 2013), addressing basic needs (BasicNeeds-Ghana), self-help groups and peer support (Cohen et al. 2012; Puschner et al. 2019). In the ongoing SCOPE project (Ethiopia), we are developing a toolkit for early identification of people who are homeless and have SMI (Hanlon et al. 2023).

**Table 3:**
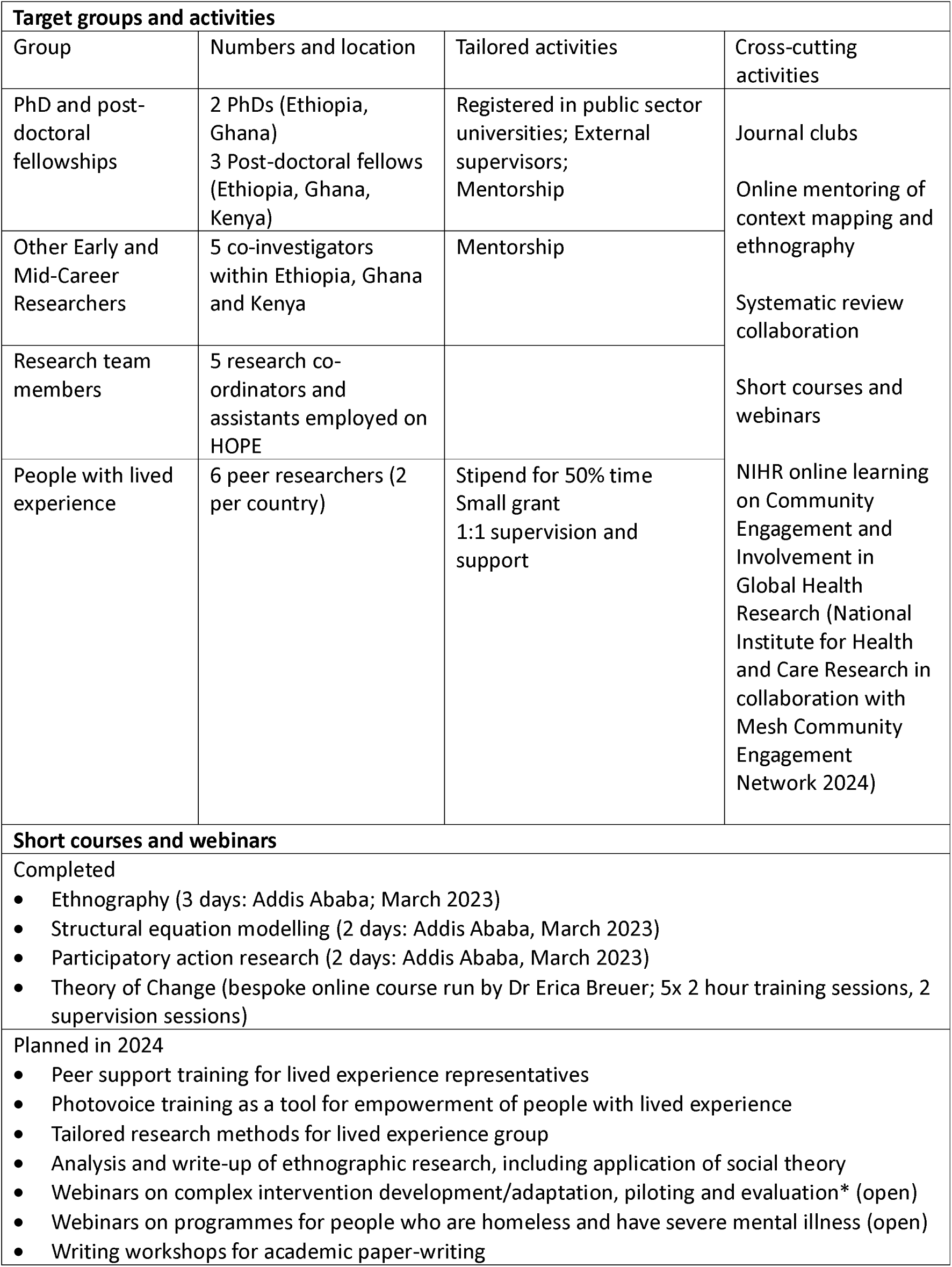
HOPE capacity strengthening activities

### Work Package 6: Lived experience, community engagement and research uptake

Mental health service user and advocacy organisations, and representatives from Ministries of Health in each partner country, contributed to the development of the HOPE proposal and were named collaborators on the grant application, in recognition of the fundamental importance of their active engagement in HOPE. A Lived Experience Advisory Group (LEAG) has been established and is meeting monthly to provide input into evolving study protocols and standard operating procedures, especially in relation to ethically sensitive aspects of the study. The LEAG then feeds back to the HOPE steering committee and investigator group monthly. We are seeking to expand the LEAG to include people who have lived experience of homelessness as well as SMI, supported by empowerment methods such as PhotoVoice (Rai et al. 2023).

Policy maker collaborators and World Health Organization representatives also join steering committee meetings on a quarterly basis and attend the annual meeting, with interim engagement within countries. In these meetings, priority is given to understanding the country level and global policy context and opportunities for HOPE to achieve impact, as well as keeping colleagues informed about, and engaged with, developing plans and emerging findings. Local community and stakeholder engagement is achieved through multi-sectoral community advisory boards and national level country management groups. Ongoing engagement and targeted messaging are key aspects of our research uptake strategy, focused on achieving high levels of ownership and the potential for sustainable impacts.

## Discussion: expected outcomes

The HOPE NIHR Global Health Research Group will speak directly to the United Nations Sustainable Development Goal imperative that ‘no one should be left behind’ in development efforts (Patel et al. 2018), seeking to address the systematic exclusion of people who are homeless and have SMI from key services and societal opportunities. While the imperative for social inclusion of people who are homeless, including those with SMI, is emphasised by the United Nations (United Nations Secretary General 2023), governments and implementing organisations are hampered by a lack of compelling and fit-for-purpose evidence; and there is a particular evidence-gap in LLMICs. In HOPE, we will generate evidence on rights-based, contextually relevant, effective and scalable interventions for people who are homeless and have SMI.

## Data Availability

All data produced in the present work are contained in the manuscript.

## Supplementary File 1 Sample size calculation for cross-sectional study

Assuming a prevalence of SMI up to 50% [1], for precision of =/-5% around the prevalence estimate, a sample of 400 people who are homeless is required per site (using a single-proportion formula). This sample size is feasible in all sites. To mitigate diagnostic accuracy issues in the field, we need to recruit more than 400 study participants. Furthermore, to look for distinct profiles of unmet needs, latent class analysis (see below) typically requires large sample sizes to ensure model convergence and proper estimation (n = 400-1000 according to empirical and simulation studies)[2]. We anticipate that in Addis Ababa, Tamale and Nairobi it will be feasible to recruit n=750 per site.

To evaluate the diagnostic accuracy (sensitivity and specificity) of community screening for people with psychosis. We assume that mental health professional assessment with SCAN (the semi-structured diagnostic assessment) is the gold standard. If prevalence of severe mental illness in people experiencing homelessness = 0.55, and

- expected sensitivity = 0.65 (this is the proportion of true positives correctly identified by community nurses)
- expected specificity = 0.95 (this is the proportion of true negatives correctly identified by community nurses)

With an expected dropout rate = 0.20, a sample of 199 would be required to estimate with 5% precision

- empirical sensitivity values that range from 0.55 - 0.75
- empirical specificity values that range from 0.85 - 1.00

Given 200 individuals who screened positive and 150 who screened negative, the target sample size for stage 2 screening is 350. This thus meets the stipulated sample size requirements. Sample size calculated using https://wnarifin.github.io/ssc/sssnsp.html

## 2 Introduction

This SOP builds on the CDT-Africa (Centre for Innovative Drug Development and Therapeutic Trials for Africa, Addis Ababa University) Safeguarding Policy. In this SOP we focus on specific procedures that are relevant to safeguarding within the HOPE project in Ethiopia.

In HOPE we will be working with people who are homeless, including those who have severe mental health conditions (MHCs). We will have contact with these individuals while on the streets and at the places where they seek help, including religious institutions, healing sites, health facilities, social care facilities and non-governmental organisations. People who are homeless are at increased risk of several types of harms and are considered a vulnerable group because of social exclusion and poverty. Those who are homeless and additionally have severe MHCs are at greater risk of harms.

In the HOPE project we have a responsibility to protect people who are homeless, with or without a severe MHC, from being exposed to harm during contacts with our project staff. We also have a responsibility to respond to any harms or safeguarding concerns that we identify in a timely and robust manner.

This Standardised Operating Procedure is based on previous approaches used when working with people with severe MHCs in the community in Ethiopia. Those ways of working were developed in collaboration with multi-sectoral Community Advisory Boards and members of the Mental Health Service User Association of Ethiopia. This draft SOP will be finalised with input from the HOPE Lived Experience Advisory Group and the Community Advisory Board in Ethiopia. We will include safeguarding as a standing item on the agenda of the twice-yearly Community Advisory Board meetings.

## 3 Purpose

This standard operating procedure (SOP) describes the process of avoiding, detecting and responding to harms and safeguarding concerns related to people who are homeless, including those who have severe MHCs, in the HOPE project.

## 4 Who is this SOP for?

This SOP applies to the following people:

- all HOPE investigators
- HOPE research staff (including the project manager, research co-ordinators, research assistants, field supervisors, peer researchers, clinicians involved in research assessments or interventions, data collectors)
- students linked to HOPE
- health extension workers, health extension worker supervisors, health workers in facilities and other professionals and community members (‘community supporters’) who are involved in the identification, referral and interventions for people who are homeless, with or without severe MHCs, as part of HOPE project activities.
- community advisory board members

## 5 Responsibility

- Everyone is responsible for working in a way that protects people who are homeless (with or without severe MHCs) against harms, conducting themselves in a way that respects their rights, and reporting any concerns about the behaviour of others promptly.
- Community supporters and community-based professionals (e.g., health extension workers) are responsible for detecting harms and safeguarding concerns, taking any necessary immediate measures and linking with the HOPE team for further actions.
- HOPE research staff working in the field are responsible for detecting and reporting harms and taking the appropriate measures, including when harms may have been perpetrated by other project staff members (whistle-blowing).
- A designated mental health focal person will support field staff in an immediate response to safeguarding concerns related to mental health, facilitate engagement with mental health care and following up to make sure that harms have been addressed to best of our ability.
- The project co-ordinator is responsible for co-ordinating a prompt response to safeguarding concerns, working with members of the research team to document any harms, concerns and actions taken (including for Serious Adverse Event reporting), and for promptly informing and involving the mental health focal person and Principal Investigators.
- Community advisory board members will promptly report any potential safeguarding concerns that they become aware of to the HOPE project team or safeguarding focal person at CDT-Africa and work closely with the HOPE team in addressing safeguarding concerns, depending on their specific area of expertise.

## 6 Abbreviations

HEW: Health Extension Worker

HOPE: Project on Homelessness and Mental Health in Africa

MHC: Mental Health Condition

PI: Principal Investigator

SAE: Serious Adverse Event

SOP: Standard Operating Procedure

## 7 SOPs for protecting against harms due to the HOPE PROJECT

### 7.1. Potential harms due to the HOPE project

Harms due to the project may occur through:

(1) The actions of project staff that exploit the vulnerable status of a person who is homeless, including those with a severe MHC.
(2) Over-riding autonomy by inclusion of a person who is homeless and lacks decision-making capacity within the project.
(3) Increased stigma against people who are homeless and have a MHC.

### 7.2. Protecting against, and responding to, harms from actions of a project worker

- All project staff employed on HOPE, students conducting projects on HOPE and HOPE investigators directly engaged in field work will first complete police checks.
- Staff, students and investigators will be trained in the Safeguarding Policy produced by CDT-Africa, Addis Ababa University and the specific HOPE safeguarding procedures.
- The safeguarding focal person for the HOPE project is the Project Co-ordinator. There is also a focal person for safeguarding in CDT-Africa.
- No person who is homeless, with or without an MHC, will be interviewed by a data collector or researcher in a private space on their own. If the person being interviewed is a woman, there will always be a woman present.
- Safeguarding concerns about a member of project staff, student or investigator can be reported by study participants to the ethics committee or using contact details on the participant information sheet.
- While conducting the community survey, the HOPE team will revisit recruitment sites 1-2 weeks after the initial assessment, giving people an opportunity to express any concerns directly.
- The community advisory board (CAB) will be an additional mechanism through which the project may hear about any concerns about project staff, investigators or students. The CAB should report any concerns directly to the HOPE safeguarding focal person or the CDT-Africa safeguarding focal person.
- If a HOPE project staff member, investigator or student observes concerning behaviour in another project staff member, investigator or student, they should immediately report this to the HOPE-Ethiopia PIs (Professor Abebaw Fekadu or Dr Ruth Tsigebrhan) or to the overall HOPE PIs (Professor Charlotte Hanlon or Professor Atalay Alem) or to the Safeguarding focal person for HOPE project or the CDT-Africa Safeguarding focal person. Who they report to directly may depend on who is involved in the safeguarding concern.
- The CDT-Africa Safeguarding Focal person should proactively engage with HOPE staff to provide opportunities for any concerns about project-linked people.
- If a safeguarding concern against a project staff member is reported to a PI, the PI is responsible for ensuring that the Safeguarding focal person at CDT-Africa is also informed. Following reporting, the CDT-Africa Safeguarding Policy guidance will then be followed with close input from the HOPE PIs. This includes considerations about whether the accused staff member needs to be suspended from field work, whether the police need to be involved and how the person who has been the victim of harms should be supported.

### 7.3. Protections for involvement of people with MHCs who lack decision-making capacity

In the HOPE project, people with severe MHCs who lack decision-making capacity to provide informed consent to participation in the study will be included in some aspects of the work, as approved by the Addis Ababa University College of Health Sciences Institutional Review Board (Ref 035/23/CDT; Date 19^th^ April 2023) and King’s College London Research Ethics Committee (HR/DP-22/23-34762; Date: 20^th^ March 2023). In the formative phase, this is relevant to the cross-sectional survey.

The justification for involvement of people who are homeless and have a severe MHC but lack decision-making capacity is the ethical principle of justice, as follows:

A. Inclusion of a person who lacks capacity to consent is justified on the following basis: Inclusion will only occur if the impairment of capacity is due to a mental health condition (confirmed by a mental health professional).
B. Research of equal effectiveness could not be carried out if confined to participants with capacity. In the previous survey of people who are homeless conducted by our team in Ethiopia, 28/89 people who were homeless and had a severe MHC lacked capacity to consent. Exclusion of this group (31% of the target population) would substantially undermine the effectiveness of the research because the purpose is to understand unmet needs to develop interventions. Exclusion of those who are most unwell would mean that those with greatest need would be least likely to benefit from future interventions.
C. The research has potential to benefit the participant directly because we will arrange linkage to support services or clinical care as needed and in line with local laws, policies and guidance. The research will also provide knowledge of the unmet needs of people who are homeless and have severe MHC to inform the development of interventions that will be implemented and evaluated in the same setting during the HOPE project.

To minimise harms from over-riding individual autonomy, the assessments have been designed to be minimally burdensome (abbreviated clinical assessment, restricting self-report to the essential information), associated with minimal risk as there will be no invasive procedures (only observation or self-report questionnaires), and will not interfere significantly with privacy (assessments will take place in a private location, with chaperone as needed) or freedom (the person is free to walk away and any expression of refusal to participate will lead to their exclusion from the project).

Procedures and protections for involvement of participants who lack capacity to consent to participation are summarised in Flowcharts 1 and 2.

Key points are elaborated below:

1. **Capacity to consent:** Assessment of capacity to consent will be undertaken by trained mental health professionals using an approach used previously in Ethiopia^1^ and requiring documentation of the key components of decision-making capacity: ability to understand information, retain it, weigh up the information and communicate a decision.
2. **Refusal:** No person communicating refusal or resistance to involvement in the study will be included.
3. **Maximising capacity to consent:** Through careful training of data collectors, we will seek to maximise an individual’s capacity to consent. Efforts will be made to explain the points included within the information sheet and allowing sufficient time for the person to ask questions.
4. **Trusted or independent individual to provide permission:**
  a. We will seek to ask the person who lacks capacity if there is a trusted individual that they would like to nominate who knows their wishes.
  b. If this is not possible, we will seek to identify whether there is an individual who is supporting the person who lacks capacity. This could be a member of a local community-based organisation or member of the public. This individual should not be a law enforcement professional or a health professional who has a clinician-patient relationship with the person. We will seek to understand how well the ‘supporter’ knows the person who lacks capacity and will only involve consultees who appear to have a good understanding of the person and their preferences. If there have only been a few brief contacts, they would not be considered as a consultee.
  c. If such a person is not available, an independent consultee would consider the individual’s circumstances and determine whether there is any evidence that the person does not want to be involved in the study and seek to identify what the person’s decision would have been based on any available information, including discussion with the person. That independent consultee will be a person from the mental health service user association collaborating with HOPE or another disability rights organisation. They will be paid for their time but not in relation to the number of people recruited.
  d. Many persons who are homeless and who have severe MHCs have lost contact with their family and some may have difficult or abusive relationships with their family (sometimes the reason that they have become homeless). We will not, therefore, seek to contact family members about participation in the study unless the person themselves wishes this and can provide their contact details.
  e. Our approach to identifying an individual to provide permission will be in keeping with the UN Convention on the Rights of Persons with Disability principles of (1) ’Will and preference’; i.e. that any nominated person is expected to arrive at a conclusion about what the person would want based on the communication they can have with them (if independent) and their past knowledge and relationship with them if a known person; and (2) ’Supportive decision-making’; i.e. that the nominated person is not seeking to arrive at just a common-sense or even ’best interest’ conclusion, but to know what the person’s decision would be so that the person’s preference is considered, whether or not they are deemed to have full capacity.
5. **Re-assessment of capacity and study withdrawal:** The person will be free to withdraw at any time. We will seek to re-assess capacity to consent and to provide opportunities for people who are included in the study to withdraw by re-visiting the recruitment area 1-2 weeks after recruitment.

For all other aspects of the HOPE formative work where consent is being obtained from a person with severe MHC, we will assess capacity at the point of recruitment and again before commencing the interview or other research activity. If during the course of the activity/interview it appears that the person does in fact not have capacity or is unable to understand the purpose of the interview/activity then we will stop the interview/activity and we will not use the data collected to that point. If the participant may regain capacity within the timescale of data collection, then we will invite them again to participate and re-assess capacity to provide informed consent. For the observational part of the ethnography, we will seek permission from gatekeepers, including police and city administration, and not from individuals.

### 7.4. Protecting against increasing mental health-related stigma and exclusion

We will seek to minimise the risk that the project could inadvertently increase stigma against people who are homeless and have a severe MHC as follows:

1. Training of all study investigators, staff and linked students on maintaining the dignity and human rights of people who are homeless, including those who also have a severe MHC.
2. Adequate supervision of project-linked people working in the field to ensure that their interactions with people who are homeless and have severe MHCs are respectful.
3. Ensuring privacy when interviews and assessments are being undertaken, meaning that interviews should not be overheard by others and, where possible, undertaken in a private space where the individual and interviewer are not visible to others (with chaperones of the same gender). This will, in part, depend on the preference of the individual as some participants may prefer to stay in a public space.
4. Ensuring confidentiality of all data obtained from participants so that details about mental health conditions do not become known to people outside the research team.
5. Working with the Community Advisory Board to address stigma as part of the HOPE activities.

## 8 SOPs for safeguarding concerns identified by HOPE PROJECT

### 8.1 Definitions of potential harms

We have identified the following potential harms that could be experienced by people who are homeless, including those with severe MHCs, and require a safeguarding approach from the HOPE project. See Table 1 for the potential safeguarding concerns and their definitions.

**Table 1:** Definition of potential harms requiring a safeguarding response

|  | Potential harm | Definition |
| --- | --- | --- |
| 1 | Chaining, restraint or seclusion | When a person is deprived of their liberty (freedom) in some way; for example, by being chained or tied up or locked in a room or in some other way restrained. |
| 2 | Suicidal behaviour or self-harm | An attempt by an individual to end their life (attempts to die by suicide) or harm themselves non-accidentally even if there is no intent to die. |
| 3 | Sexual exploitation, abuse or harassment | <u>Sexual abuse</u> is an actual or threatened physical intrusion of a sexual nature, whether by force or under unequal or coercive conditions. Sexual abuse includes sexually touching a person without their consent (e.g. groping or forced kissing). Rape is a form of sexual assault involving penetration of the vagina, anus or mouth.<br><u>Sexual exploitation</u> is any actual or attempted abuse of a position of vulnerability (e.g. mental illness), differential power or trust for sexual purposes. Sexual exploitation includes child sexual exploitation, exploitation of person with psychosis to be involved in sexual activities, project workers exchanging sex for money or goods or services.<br><u>Sexual harassment</u> includes sexually offensive comments, unwanted sexual advances, requesting sexual favours, gesturing or making sexual remarks about someone's body, clothing or appearance. |
| 4 | Physical abuse | Slapping, smacking, hitting, punching, kicking or spitting at someone or beating or harming the person with an implement (e.g. stick, knife). |
| 5 | Severe physical neglect or ill-health | Due to lack of basic needs and access to care, the person's health and/or nutritional status are at severe risk without intervention. |
| 6 | Neglect of children who live on the streets | Failure or absence of a caregiver to provide needed food, shelter, clothing, medical care, or supervision to the degree that a child's health (including nutritional status), safety, and well-being are at risk. |
| 7 | Violence or aggressive behaviour towards others | In the context of inadequate treatment and support, a minority of persons with severe MHCs may become violent or aggressive to others. |
| 8 | Coercion to take treatments or engage with services | Person is forcibly given treatment against their will. This could be medication or religious or traditional treatments or forcible removal to a service. |
| 9 | People trafficking | The children of people who are homeless, including those with severe mental health conditions, could be vulnerable to being trafficked by criminals. |
| 10 | Drug dealing or criminal activities | The person may be involved in criminal activities or dealing drugs. |
| 11 | Labour exploitation | The person may be exploited for their labour without remuneration or safe working conditions. |

### 8.2 Identification of harms

Harms against people who are homeless, with or without MHCs, may be identified through direct observation or through disclosures from the person themselves or reports from others. The most relevant means of identification of these harms in the context of HOPE are as follows:

| Type of harm | Identification |
| --- | --- |
| Restraint, chaining or seclusion | Observed in a community setting, religious or traditional healer setting, social care facility or in health facilities. |
| Suicidal behaviour or self-harm | Reported by the person who is homeless (whether or not they have a diagnosis of a severe MHC) in the context of research activities, or during community case detection or during help-seeking at healing sites, social care facilities or in health facilities. |
| Sexual abuse, harassment or exploitation; Physical abuse; Severe physical health or neglect; Labour exploitation | Directly observed or reported by the person or local community members in the context of research activities, or during community case detection or during help-seeking at healing sites, social care facilities or in health facilities. |
| Violence or aggression towards others; involvement in drug dealing or criminal activities | Directly observed or reported by local community members in the context of research activities, or during community case detection or during help-seeking at healing sites or in health facilities. |
| Neglect of children who live on the streets; People Trafficking | Directly observed or reported by the person or local community members in the context of research activities, or during community case detection. |
| Coercion to take treatment or engage with services | Directly observed or reported by the person with severe MHC in the context of research activities or during help-seeking at healing sites or in health facilities. |

### 8.3 Actions to be taken upon identification of safeguarding concerns

Flowcharts summarising the required response are presented in the Appendix.

The researchers will provide all participants with contact details for support and advocacy services as part of the recruitment process.

#### 8.3.1 Chaining, restraint or seclusion in public settings

See flowchart 1. Note, for restraint or seclusion taking place on the premises of an organisation, refer to flowchart 6.

- Chaining, restraint or seclusion should be considered an emergency requiring action on the same day.
- The project-linked person will seek to understand why the person has been chained, restrained or secluded, and by whom, and explain the need for mental health care.
- The project-linked person will communicate with the project mental health focal person to plan how and where mental health care can be accessed.
- If the person who is chained, restrained or secluded <u>is willing to attend</u> for mental health care, the project-linked person will work with the health extension worker and/or community police to convey the person to that facility.
- If the person who is chained, restrained or secluded <u>refuses to attend</u> for mental health care, the project-linked person will work with the community police to convey the individual to mental health care.
- If a hospital admission is needed, the project mental health focal person will report this as a Serious Adverse Event.
- The project mental health focal person will co-ordinate with HOPE team and seek to ensure that the person has received care, is no longer restrained in the community and has ongoing mental health support.

#### 8.3.2 Sexual abuse, exploitation or harassment

See flowchart 2.

- The project-linked person will first and foremost be supportive, take the allegations seriously and ensure privacy.
- If there is any concern that the situation is not safe, e.g. because a perpetrator of sexual assault is still in the vicinity, the project-linked person will re-locate to a safe place, with the affected individual if possible. A safe place means anywhere out of the vicinity or could be a government health facility. If that is not possible, the interview will stop and the project-linked person will inform the HOPE project safeguarding focal person immediately.
- The project-linked person will not force the person to speak about their experiences. They will be trained in principles of Psychological First Aid and will use these skills if the person is distressed. The project-linked person will ask about potential supports for the person and seek to mobilise support, with the person’s permission.
- If the person reports sexual abuse, the project-linked person will encourage and support attendance at Ghandi hospital one-stop service for further assessment on the same day.
- The project-linked person will explain that they will need to speak to their seniors but that information will be kept confidential.
- Information about potential supports and places to go for help will be given verbally. If willing, the person will be supported to access these supports e.g. with provision of transport and being accompanied by a project worker.
- The HOPE safeguarding focal person should be informed on the same day and will communicate with the PIs to determine whether a sexual assault has taken place and whether a Serious Adverse Event (SAE) should be reported.
- The project mental health focal person should also be informed so that any necessary mental health support can be arranged, regardless of whether or not the person has a severe MHC.
- If a severe MHC is suspected, the project-linked person will liaise with the project mental health focal person so that an urgent assessment of decision-making capacity can be made.
- If the person wishes to report the incident to the appropriate authorities (e.g. Women and Social Affairs; police), the project will facilitate (e.g. with transport, accompanying the person), as needed.
- If the person has no MHC or if they have a severe MHC but are assessed to have decision-making capacity, If they wish not to report the incident, this will be respected.
- If the person has a severe MHC and lacks decision-making capacity and does not wish to report the incident, the PIs will be informed. An urgent risk assessment will be conducted to determine whether to report the incident against the person’s wishes. The decision will be recorded with justification.

#### 8.3.3 Physical abuse or severe ill-health

Flowchart 3.

- If there is any concern that the situation is not safe, the project-linked person will re-locate to a safe place, with the affected individual if possible. A safe place means anywhere out of the vicinity or could be a government health facility. If that is not possible, the interview will stop and the project-linked person will inform the HOPE safeguarding focal person immediately.
- If someone is at immediate risk of serious harm of physical abuse, for example being threatened or attacked with a weapon, or if the project-linked person found that someone had been seriously injured due to physical abuse, or even killed, then the research team would inform the police in order to take urgent action and immediately report the incident to the HOPE safeguarding focal person and/or one of the HOPE PIs.
- If physical abuse is being perpetrated by the police, the project-linked worker will immediately contact the HOPE safeguarding focal person and/or one of the HOPE PIs. It is essential that the project-linked worker should prioritise their own safety. The incident will be followed up by the HOPE PIs and safeguarding officer, including a discussion about whether the police officer can or should be reported. In any case, HOPE will engage with the police via the community advisory board and directly to promote approaches that uphold the human rights of people who are homeless, including those who have a severe MHC.
- If there are serious concerns about the person’s physical health e.g. due to physical abuse, accidental injury or because of medical illness or severe nutritional deficiency, the project-linked worker will urgently contact the family health team or health extension worker to arrange access to emergency health care.
- The HOPE safeguarding focal person should be informed on the same day and will communicate with the PIs to determine whether a Serious Adverse Event (SAE) should be reported.
- If a severe MHC is suspected, the project-linked person will liaise with the project mental health focal person so that an urgent assessment of decision-making capacity can be made.
- If the person wishes to report the incident to the appropriate authorities, the project will facilitate (e.g. with transport, accompanying the person), as needed.
- If the person has no MHC or if they have a severe MHC but are assessed to have decision-making capacity, if they refuse health care or decline to report the incident, this decision will be respected. In that case, the person will be provided with information about how and where they can seek help should they want it.
- If the person has a severe MHC and lacks decision-making capacity and does not wish to access healthcare or report the incident, the PIs will be informed. An urgent risk assessment will be conducted to determine whether to override person’s wishes. The decision will be recorded with justification.

#### 8.3.4 Suicidal behaviour or self-harm

Flowchart 4.

- The project-linked person will first and foremost be supportive and respectful towards the person.
- The project-linked person will stay with the person and remove potential means of suicide in the vicinity.
- They will contact the family health team or health extension worker and support urgent attendance of the person for a mental health assessment in the health centre or other health facility.
- The project-linked person will inform the HOPE safeguarding focal person who will complete reporting for an Serious Adverse Event (SAE) if relevant.
- The project-linked person will contact the HOPE mental health focal person to plan how and where mental health care can be accessed.
- If the person refuses mental health care, the HOPE mental health focal person will arrange for urgent assessment of decision-making capacity, suicide risk and mental health needs.
- If the person is assessed as having a high suicide risk and lacks decision-making capacity and refuses mental health care, project staff will co-ordinate with the community police to convey to mental health care.

#### 8.3.5 Violent or aggressive behaviour

Flowchart 5.

- If there is any concern that the situation is not safe, the project-linked person will re-locate to a safe place, the interview will stop and the project-linked person will inform the HOPE safeguarding focal person immediately.
- The project-linked person should prioritise their own safety.
- If it is safe to do so, they should speak calmly and respectfully to the person throughout and encourage others to do the same.
- If a member of the public is injured, they should support them to access health care by arranging transport.
- If the person appears to have a mental health condition, work with community police to arrange urgent transfer to mental health services.
- If there is no apparent mental health condition, inform the community police so that they can take appropriate action.
- The project-linked person will then report to the HOPE safeguarding focal person who will determine whether a Serious Adverse Event has occurred i.e., if there is violent behaviour causing injury or arrest/imprisonment.
- If the person has a mental health condition, the HOPE mental health focal person will be informed. The HOPE mental health focal person will help to co-ordinate access to mental health services and will report a Serious Adverse Event if hospitalisation is required.

#### 8.3.6 Coercive treatment, restraint or abuse on the premises of facilities or healing sites

Flowchart 6.

- Any of the above safeguarding concerns could take place on the premises of an organisation; for example, a health facility, social care facility, traditional or religious healing site, church, mosque or police station.
- In such situations, if there is any concern that the situation is not safe, the person working with the HOPE project should re-locate to a safe place and inform the HOPE safeguarding focal person immediately.
- If there is any immediate and serious threat to the person, inform the police immediately. However, if the police are the perpetrators, the project-linked person should first prioritise their own safety. They should then inform the HOPE safeguarding focal person and/or PIs immediately.
  - The incident will be followed up by the HOPE PIs and safeguarding focal person, including a discussion about whether the police officer can or should be reported.
  - Plans will be made to discuss with the multi-sectoral Community Advisory Board in each study area to plan the best way to tackle such practices and incidents on an ongoing basis. Usually this will be on a 6-monthly basis, but if needed the senior research team will convene a sub-committee to address a specific issue.
- If action is safe, the project-linked person will follow the relevant flow chart, depending on the nature of the safeguarding concern.
- The HOPE safeguarding focal person should be contacted on the same day and will determine whether an SAE needs to be reported.

#### 8.3.7 People Trafficking or involvement in Drug Dealing or Criminal Activities

Any concerns about possible people trafficking activities, drug dealing or criminal activities that put others at immediate risk will be reported directly by project-linked person to the HOPE safeguarding focal person and from there to the PIs. This senior management team will then report any concerns to the appropriate authorities.

#### 8.3.8 Neglect of children living on the street

Any concerns about serious neglect of children (under 15 years of age) living on the street will be reported to the Women and Social Affairs Office and local health team.

## 9 APPENDIX: FLOW CHARTS FOR RESPONSES TO SAFEGUARDING CONCERNS

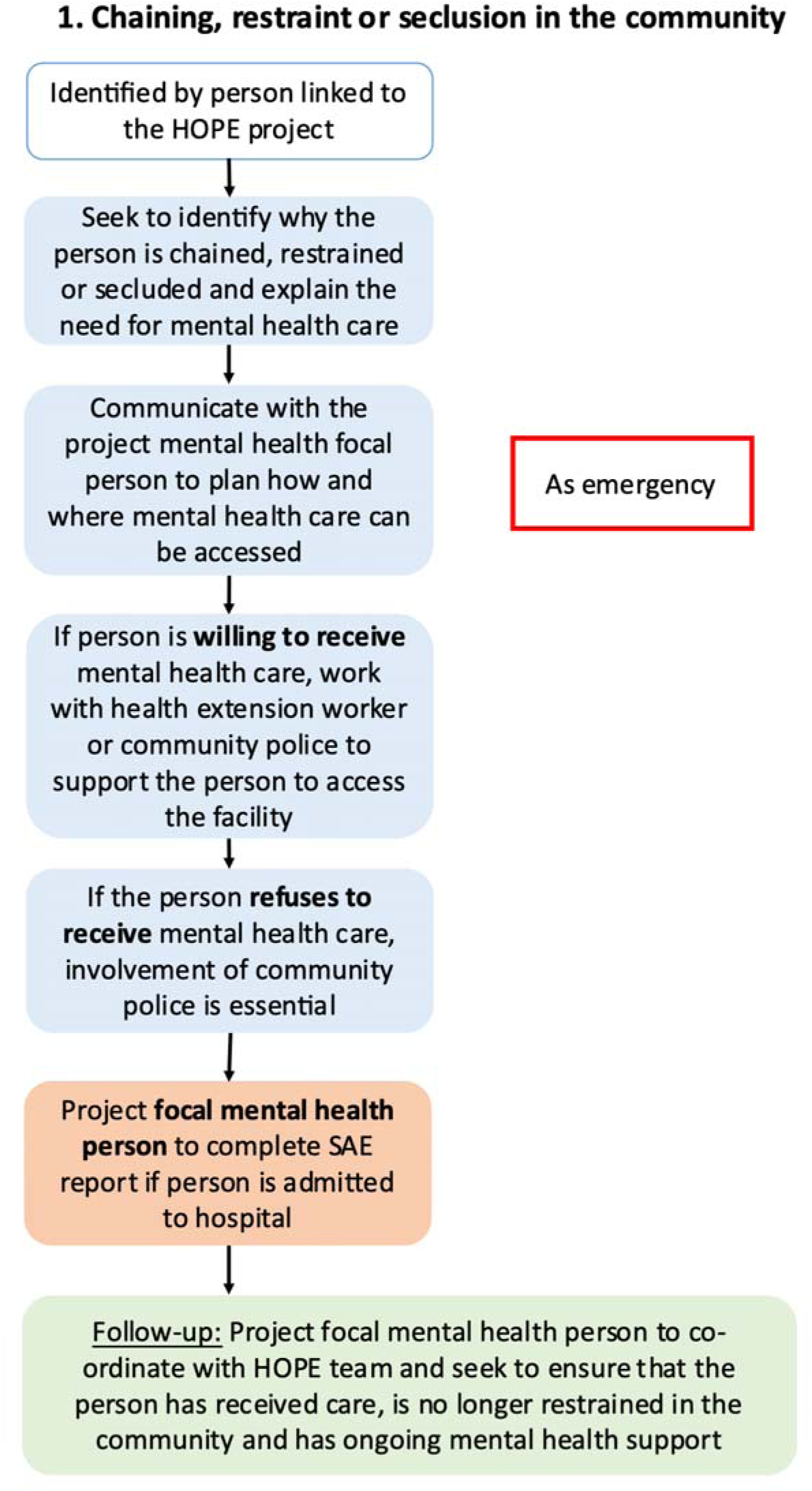

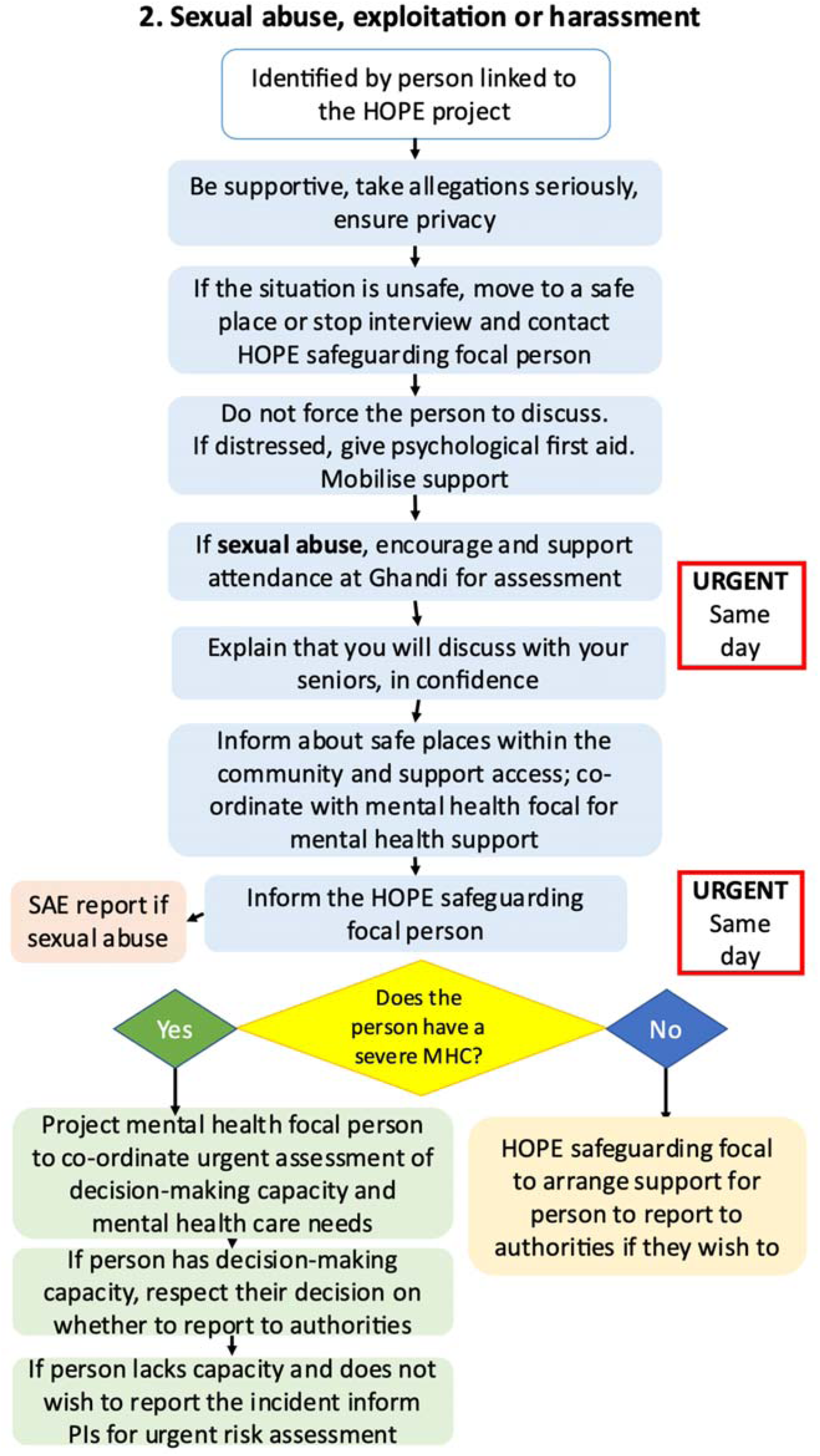

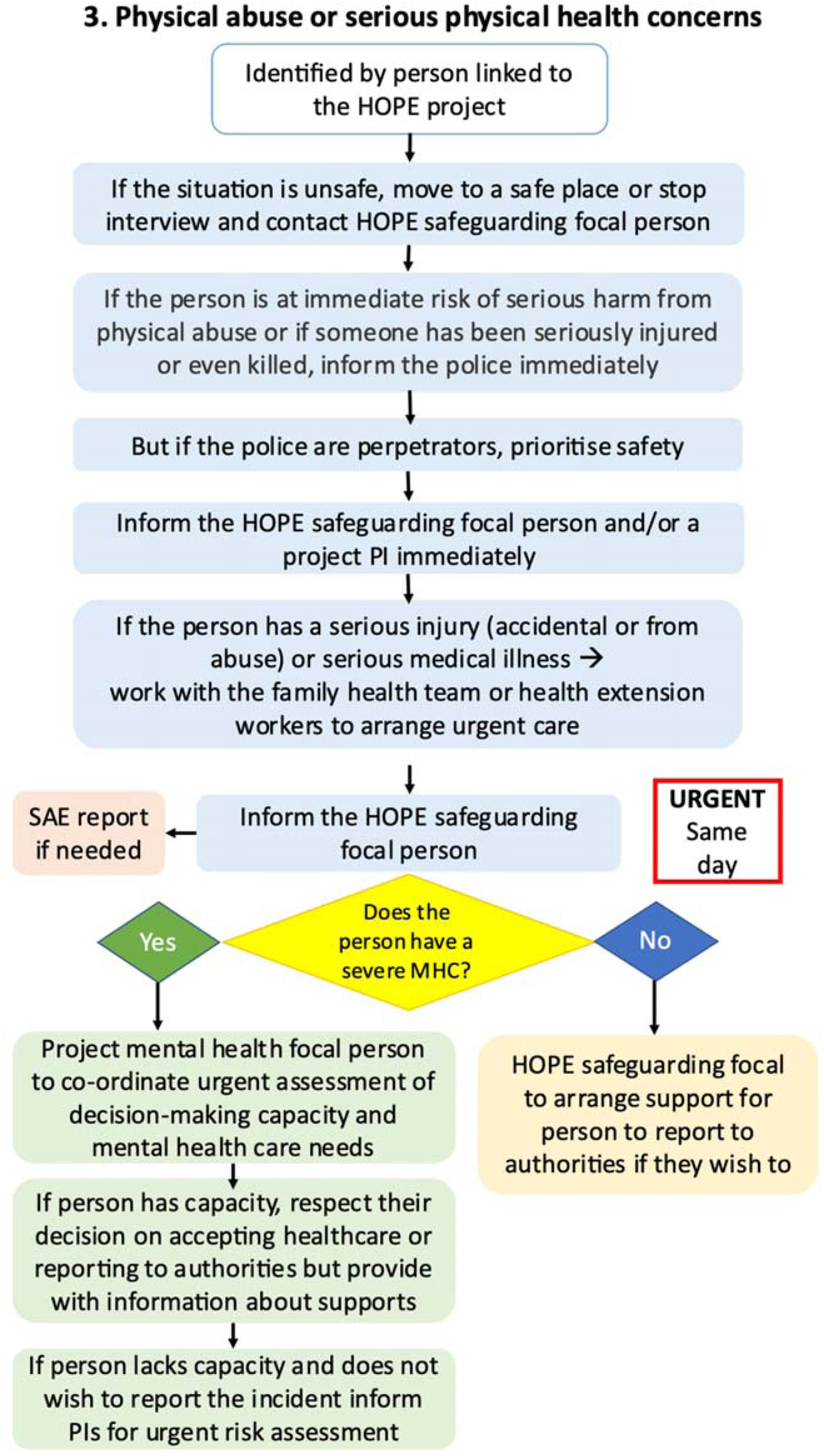

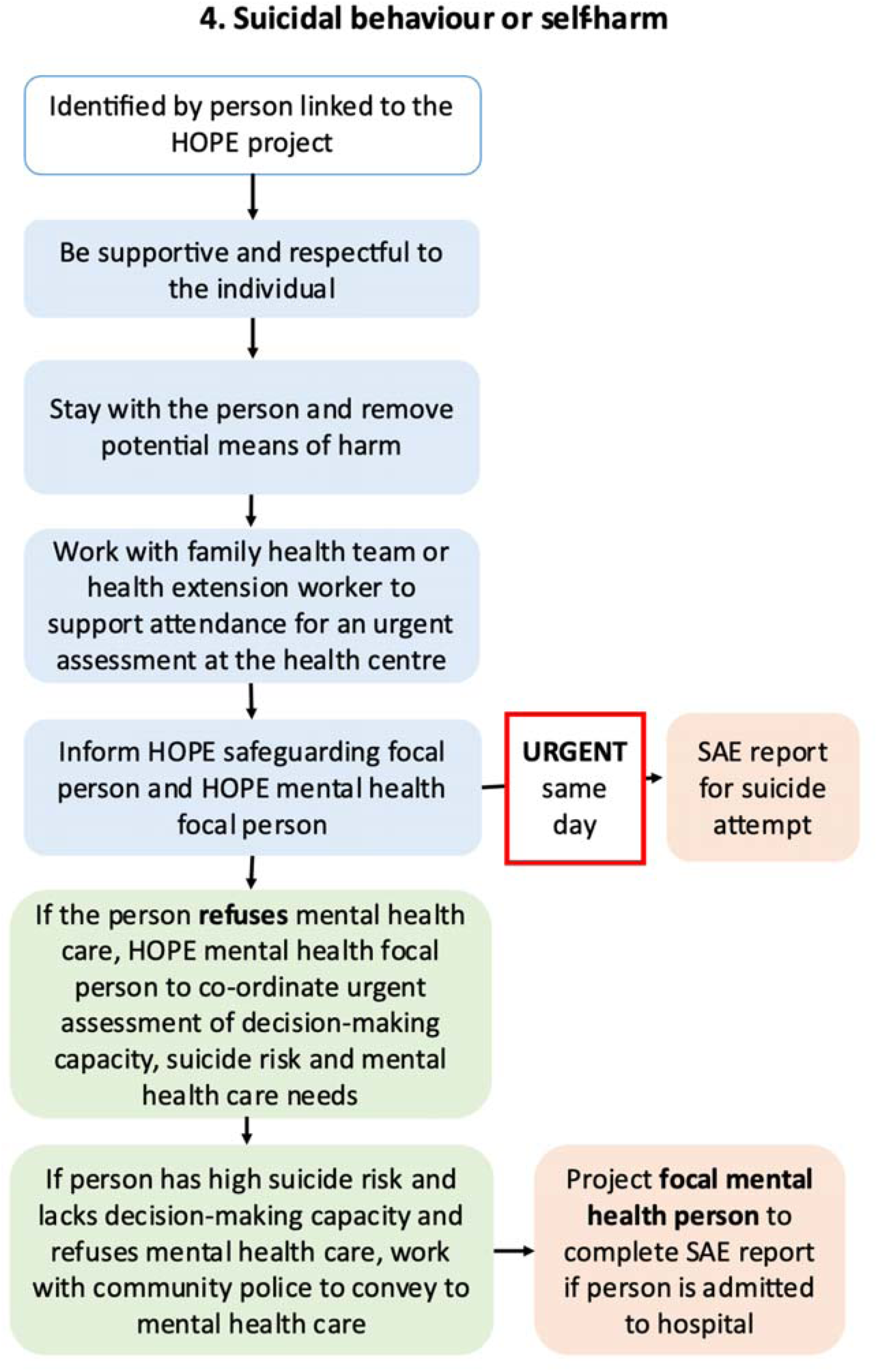

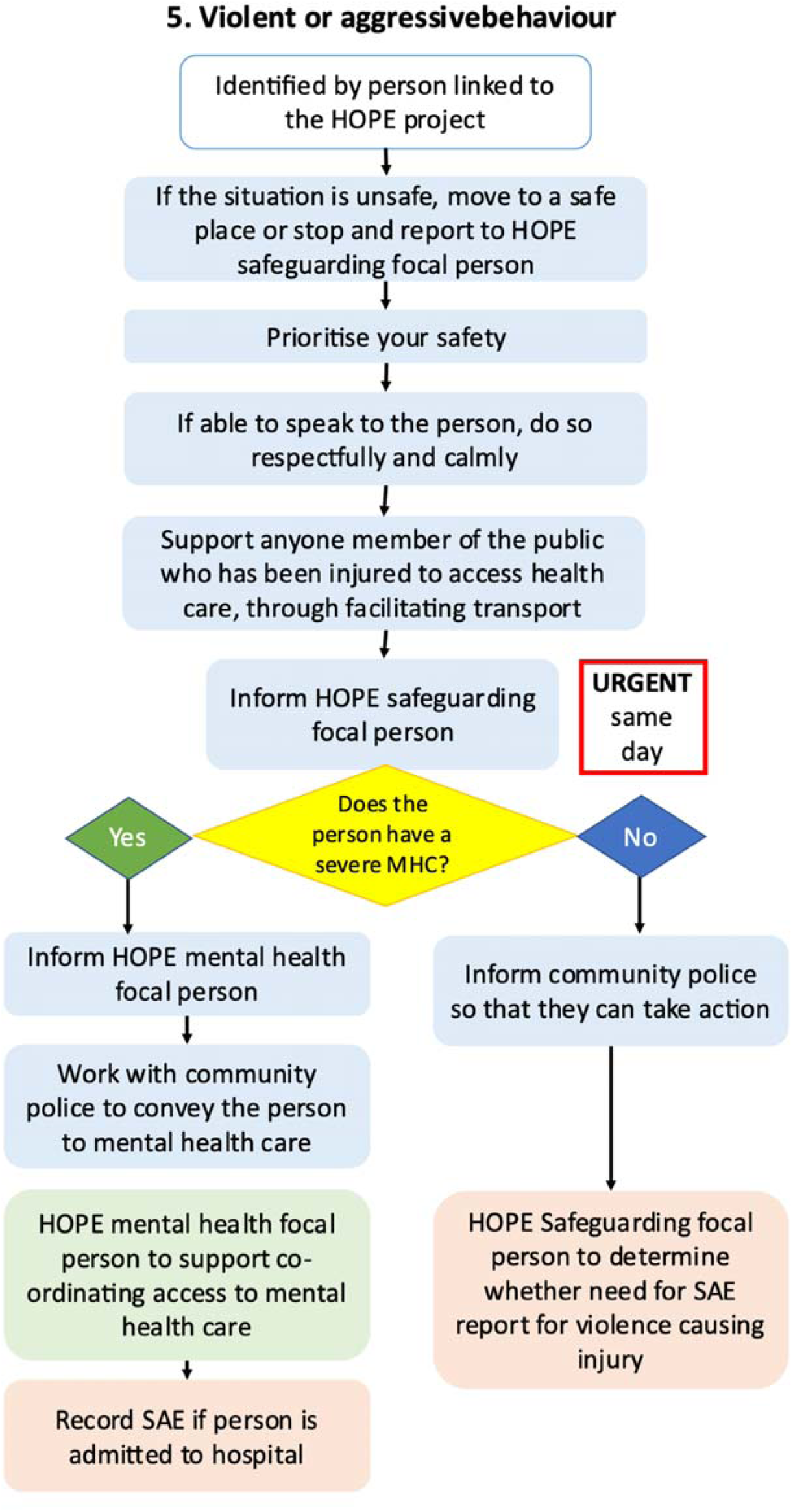

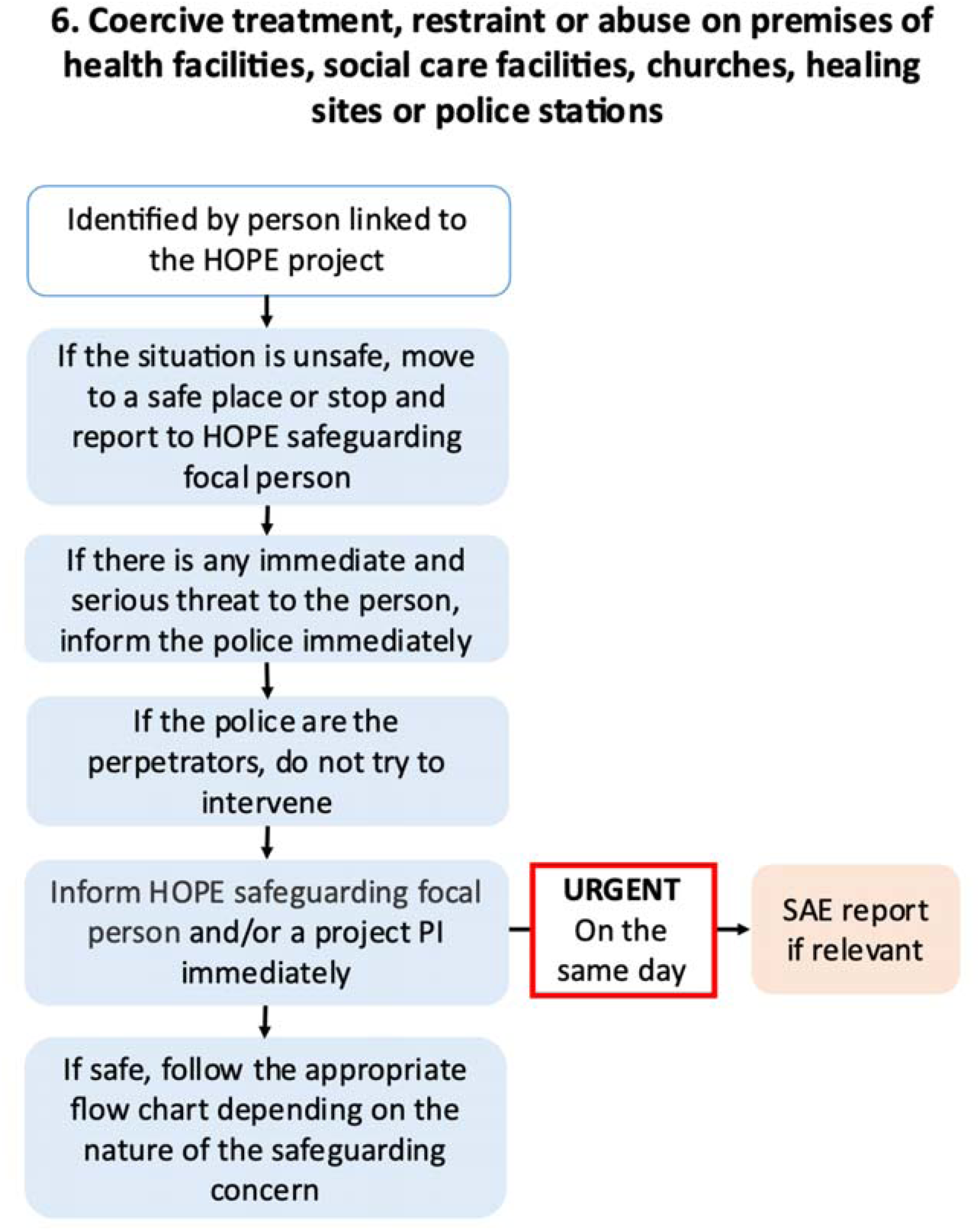

## Footnotes

1 Hanlon C, Medhin G, Dewey ME, et al. Efficacy and cost-effectiveness of task-shared care for people with severe mental disorders in Ethiopia (TaSCS): a single-blind, randomised, controlled, phase 3 non-inferiority trial. Lancet Psychiatry 2022;9(1):59-71. doi: 10.1016/s2215-0366(21)00384-9

